# Toward comprehensive multimodal health monitoring in heat: A digital biomarker exploration study

**DOI:** 10.64898/2026.09.17.26363299

**Authors:** Léa Schüpfer, Carmela Niederberger, Vanessa Matos Gonçalves, Jörg Goldhahn, Noé Brasier

## Abstract

**Background:** As climate is significantly changing, the need for effective physiological health monitoring during heat exposure becomes increasingly urgent. Population-based indices provide valuable environmental insights but fail to account for individual variability in heat response. A person-centered approach to heat strain assessment remains lacking. Digital biomarkers offer a promising avenue for individualized, continuous, and lab-independent health monitoring in real time. Here, we explored digital biomarker candidates for heat health monitoring.

**Methods:** We conducted a single-center, prospective, cross-sectional, cross-over, observational, explorative study to identify digital biomarker candidates for health management during heat exposure. 20 participants were exposed to various personalized heat stressors. Vital parameters, such as heart rate, core and close body temperature were assessed using wearable sensors, and the local sweat rate was determined using technical absorbents. A Random Forest model was built to distinguish between ambient temperature, relative humidity, and physical exertion through digital biomarker candidates.

**Results:** The Random Forrest model distinguished between the heat stressors with 94.7% accuracy. Key contributing variables differed by stressor type, highlighting the value of multimodal sensing including physiological and environmental factors. The variables distribution for each stressor revealed varying patterns: predictors for ambient temperature were primarily environmental measures, for exertion primarily physiological assessments, and relative humidity stressors physiological and environmental measures.

**Discussion:** Our findings reveal the potential of digital biomarkers to differentiate heat exposure origins and support the development of comprehensive automated, lab-independent, and person-centered health monitoring strategies. Further, this work showed the promise of integrating physiological as well as environmental measures to enable comprehensive health monitoring during heat exposure. Further testing is needed in more complex and real-life settings.

## Introduction

Extreme weather events pose an increasing threat to human health and safety, leading to elevated rates of accidents, illness, and mortality^1^. With global temperatures rising, climate change intensifies heat stress, increasing the risk of impeding physiological processes and their respective perception of comfort^2^. Heat stress can result from a variety of factors, including high ambient temperatures, elevated humidity, increased ambient solar radiation, along with actively generated heat from physical exertion, and restricted heat dissipation when wearing protective clothing^3^. Thus, in a complex real-world setting, heat stress is often caused by multiple factors^4^ at the same time. While vulnerable groups such as people of young/advanced age, chronically ill, and individuals unable to care for themselves are particularly at risk when exposed to ambient heat, healthy and young people are at risk during extensive physical exertion, such as during physical work ^4^. In response to heat stress, the human body activates physiological strategies to maintain thermoregulatory homeostasis and stabilize core body temperature at around 37 °C. This physiological body reaction is collectively referred to as heat strain^5^. Heat strain includes sweating, increased peripheral vasodilation, and behavioral adaptations such as reducing activity or removing clothing^5^. The magnitude and effectiveness of these responses are influenced by individual characteristics such as age, physical fitness, heat adaptation, and overall health^6^. Still, these safety mechanisms of the human body might be insufficient in various situations, putting humans at imminent and potentially significant health risk ^4^. Therefore, it is of utmost importance to monitor heat stress and identify people at risk as early as possible to prevent them from deteriorating health events.

Heat stress monitoring and management traditionally rely on both population-based measures, such as the wet-bulb-globe-temperature (WBGT) and the Universal Thermal Climate Index ^7^, to assess environmental conditions. However, a person-centered approach to assess heat strain and evaluate how heat affects an individual’s health is still missing. While environmental metrics and metabolic heat estimates are commonly used to assess heat stress and partly explain it, they have failed to fully account for the wide inter- and intra-individual variability in physiological responses of the individuum^8^. As such, personalized physiological monitoring is increasingly recognized as a critical tool for providing tailored health management and protecting against heat-related illness^8^.

Recent advancements in connectable wearable devices capable of measuring a variety of health metrics^9^ have the potential to disrupt state of the art health care provision. Wearable sensors allow for continuous and mostly non-invasive monitoring of digital biomarkers including core body temperature (CBT), skin temperature, heart rate (HR), local sweat rate (LSR) in both the lab, and in the field^10^. By incorporating these digital biomarkers into heat stress exposure management, wearable devices promise more personalized and accurate health monitoring. Historically, these sensors focus on sensing vital parameters - “classical” heat strain measurements. Even this sensing approach offers a novel way to personalize heat stress measurements, yet these devices lack sensing important context information that is crucial for meaningful medical care. To achieve this more differentiated physiological understanding, environmental information must be included together with personal vital sign sensing^11^. Data interpretation in unsupervised real-life settings is challenging as the source of heat stress can vary. A more personalized and accurate monitoring of heat stress could be achieved by not only incorporating the afore mentioned “classical” heat strain measures into heat strain management, but by also looking at the environmental circumstances. This also allows for implementing adequate counter measures as a reaction to a comprehensive understanding of human physiology in the heat.

In this study, we explored digital biomarker candidates towards continuous and automated health monitoring in the heat. For this, participants were exposed to personalized heat stress from the environment and exertion in a controlled environment to enable a fine-grained analysis of biomarkers related to the respective isolated heat stressor.

## Results

### Study participants

A total of 23 participants were enrolled in the study between November 2022 and February 2023. Three participants withdrew voluntarily, and two heat stress study visits were discontinued prematurely due to sensor failure and were therefore excluded from the analysis. Ten female and ten male participants were included in the final analysis. Their average age was 25 years (SD± 4.37). The participant flow chart of the study was published previously^12^. Seven days prior to visit 2, iButton sensors were used to measure the close body temperature (CloBT) and the close body relative humidity (CloBH) about 6 mm above the body surface. The mean CloBT was 29.89 °C (SD ± 1.89) and the CloBH was 37.08% (SD ± 7.47) (a differentiated information about participant characteristics is available in the publication from Brasier et al.^12^).

WBGT, an indicator for environmental heat stress, showed significant differences under increased ambient temperature and relative humidity conditions (both considering protective clothing), compared to exertion, which primarily reflects active heat stress (Table 2). This heat stress resulted in higher whole body sweat loss during ambient heat stress (0.16 L/m^2^ ± 0.05 L/m^2^) compared to relative humidity (mean 0.07 L/m^2^ ± 0.03 L/m^2^) as discussed earlier^12^. However, the highest whole body sweat loss was achieved during exertion (mean 0.3 L/m^2^ ± 0.07 L/m^2^), underlining the fact that monitoring the environmental heat is not enough to assess and monitor the impact of heat on the human body. While significant increases in WBGT were observed during the Stress and Stress+Clothing (“Stress+Cl”) phases for both ambient temperature and relative humidity in relation to the acclimatization (all *p* < 0.0001), no statistically significant differences were found regarding the exertion phase (all *p* > 0.66) (Table 1).

**Table 1.** Mean WBGT temperature per stress-phase pair and mean differences (confidence intervals) between acclimatization and other phases within a stressor.

| Phase | Estimate (°C) | Difference (°C) | Confidence Interval (°C) | p-value |
| --- | --- | --- | --- | --- |
| **Temperature** |  |  |  |  |
| Acclimatization | 23.24 | NA | NA | NA |
| Stress | 31.26 | 8.02 | (6.95 to 9.09) | < 0.0001 |
| Stress + Cl | 31.39 | 8.15 | (7.08 to 9.22) | < 0.0001 |
| Cool | 23.28 | 0.04 | (-1.03 to 1.11) | < 0.95 |
| **Relative Humidity** |  |  |  |  |
| Acclimatization | 23.16 | NA | NA | NA |
| Stress | 27.74 | 4.58 | (3.51 to 5.65) | < 0.0001 |
| Stress + Cl | 27.91 | 4.75 | (3.68 to 5.82) | < 0.0001 |
| Cool | 23.31 | 0.15 | (-0.92 to 1.22) | 0.78 |
| **Exertion** |  |  |  |  |
| Acclimatization | 23.29 | NA | NA | NA |
| Stress | 23.53 | 0.23 | (-0.86 to 1.33) | 0.68 |
| Stress + Cl | 23.42 | 0.13 | (-0.97 to 1.23) | 0.82 |
| Cool | 23.04 | -0.25 | (-1.35 to 0.85) | 0.66 |

### Physical health monitoring in heat

#### Core Body Temperature

During the “Stress” phases, core body temperature (CBT) generally showed an upward trend in most subjects, particularly when exposed to exertion. This increase was most prominent in Subjects 1, 7, 10, 12, 18, and 19. In several cases, CBT continued to rise during the “+Clothing” phase (Supplemental Information 1). In contrast, Subjects 3, 13, 19, 21, and 23 showed relatively stable CBT profiles throughout the whole session, with minimal fluctuations despite changes in environmental (temperature and humidity stressor) conditions. A decrease in CBT during the “Cool” phase was observed in all subjects. However, 30 minutes of cool-down was not enough time to reach initial baseline CBT, especially in the high-stress exposure visits. The highest mean values of CBT were observed during the “Heat + Clothing” phase for all stressor types and highest with exertion: ambient temperature (37.21 °C ± 0.48), relative humidity (37.22 °C ± 0.22), and exertion (37.79 °C ± 0.26) (Supplemental Information 1.2).

### Heart Rate and Heart Rate Variability

During the “Stress” phases, heart rate (HR) increased in all participants, the highest response was observed during physical exertion (Supplemental information 1.3). This effect was most pronounced in Subjects 3, 4, 10, 12, and 21, who reached HRs of up to 160 bpm during exertion. In these individuals, the rise in HR closely followed the onset of exertion and remained elevated during the “+Clothing” phase. In contrast, Subjects 2, 6, 8, 9, and 20 exhibited relatively stable HR profiles, particularly during the ambient temperature and humidity stressor phases, with only minor fluctuations observed throughout these sessions. Under ambient temperature and relative humidity conditions, HR increases were generally modest, with most values remaining below 100 bpm. Baseline HR during the acclimatization phase ranged from 65 to 85 bpm across participants. A gradual return toward baseline was observed during the “Cool” phase in most subjects (Supplemental Information 1.4). Interestingly, subjects 2, 4, 12, 15, and 20 showed a relatively flat rMSSD (Root mean square of the successive differences) profile across all phases, with only minor variation over time. In contrast, others exhibited more dynamic patterns, such as in subjects 1, 7, 8, 11, 13, and 22, with noticeable peaks and drops between phases throughout all three stressors. In several cases, rMSSD values decreased during “Stress” and “+Clothing” phases and increased during the “Cool” period (Subjects 1, 7, 8, 11, 19, and 21). This variability highlights substantial inter-individual differences in short-term HRV and dynamic autonomic responses to environmental and exertional stressors (Supplemental Information 1.5). However, in subject 20, there was almost no variability.

The SDNN (Standard Deviation of NN intervals) time series data showed a high degree of fluctuation across all phases and stressor conditions (Supplemental Information 1.6). In most participants, SDNN values varied continuously intrapersonal throughout the session, including during the “Acclimatization”, “Stress”, and “Cool” phases. Most subjects displayed frequent and irregular peaks and drops, regardless of the applied stressor. Some participants, including Subjects 2, 4, 12, and 20, had lower and more stable SDNN values overall, but still showed short-term fluctuations without a consistent pattern. Notably, during the exertion stressor (blue line), a reduction in SDNN was observed during “Stress” and “+Clothing” phases in most subjects (Supplemental information 1.6). These patterns indicate substantial inter- and intra-individual variability in SDNN throughout the measurement period. Again, subject 20 shows almost no variability in its SDNN profile.

The highest mean HR was observed under exertion, rising from 79 bpm (SD ± 14.01) during “Acclimatization” to 130 bpm (SD ± 16.95) during the “Heat + Clothing” phase. In comparison, smaller increases were seen under ambient temperature (79 bpm ±14.01 to 86 bpm ± 15.25) and relative humidity (77 bpm ±11.77 to 78.05 bpm ±12.17). Standard deviations across all phases remained relatively stable, indicating consistent variability between participants (Supplemental Information 1.4).

During exertion, a negative correlation between the HR slope and VO_2_max was identified (a 1-unit increase in VO_2_ max is associated with a decrease by 0.019 in heart rate, *p*-value = 0.045) (Figure 1). Thus, study participants who had a lower VO_2_max showed a steeper HR slope during standardized exertion compared to those with a higher VO_2_max.

**Figure 1.**
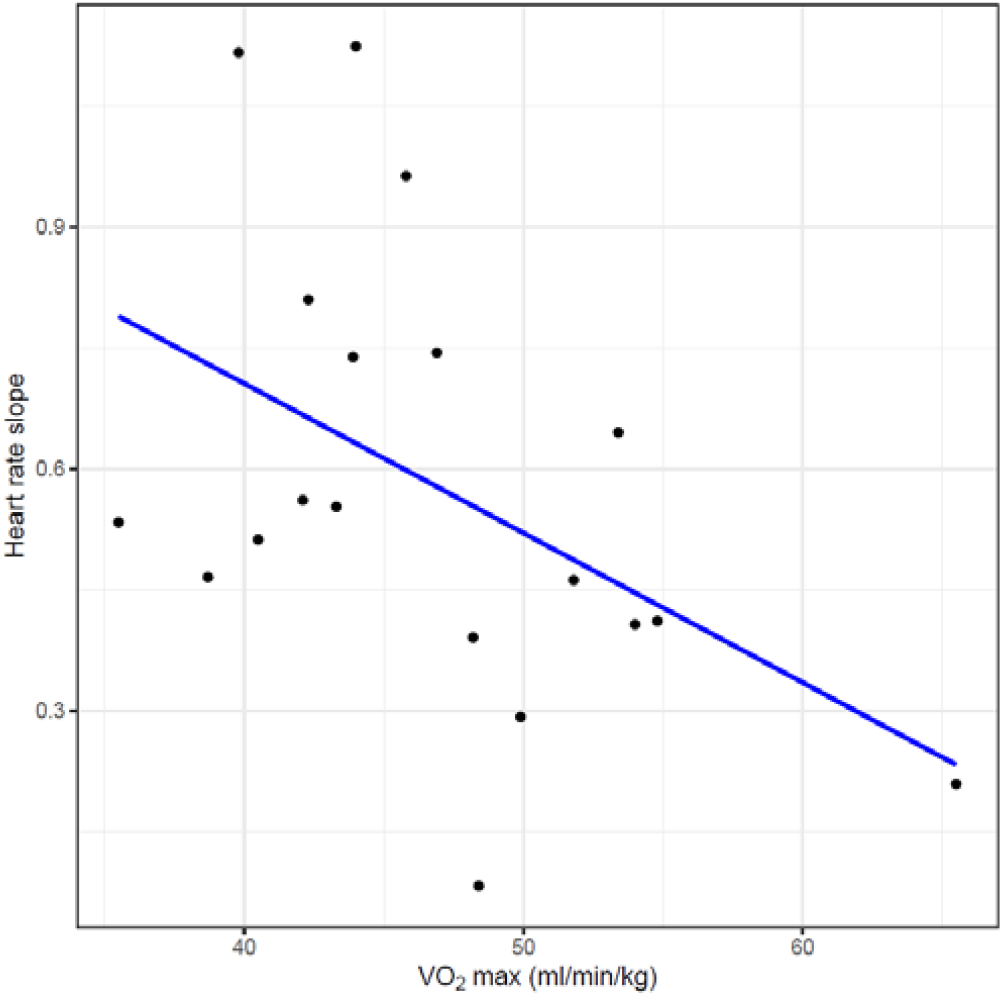
Scatterplot of heart rate slope versus VO_2_max with fit from linear model.

### Sweat rate

During the “Stress” phase, the LSR, measured using a gold-standard plaster-based method, increased in most participants. A further rise in LSR was often observed during the “+Clothing” phase (Supplemental Information 1.7). Subject 12 showed the maximum value of 3221 g/h*m^2^ between minutes 90 and 120 of the exertion visit. For most participants exposed to ambient heat stress, LSR progressively increased across the phases, reaching its peak at the end of the “+Clothing” phase, followed by a decline during the cool-down phase. However, peak values during ambient heat exposure remained lower than those observed during the exertion stress phase. During the relative humidity stressor, LSR was low and remained relatively stable across all phases in most participants, with minimal increases observed throughout the session.

Mean LSR increased across all phases for each stressor condition. The largest increase occurred under exertion, where the mean LSR rose from 25 g/h*m^2^ during acclimatization to 967 g/h*m^2^ during “Heat+Clothing”, accompanied by high inter-individual variability (SD = ±640 g/h) (Supplemental Information 1.8).

### Close body temperature and humidity

Close body thermal conditions, reflected by the temperature and relative humidity measured with the iButton sensor, worn by each participant varied consistently in response to the different stressor exposures. During exposure to ambient heat, the CloBT increased rapidly during the “Stress” phase and peaked during the “+Clothing” phase for most participants, followed by a gradual decline during the cool-down phase (Supplemental information 1.9a). Under elevated relative humidity and exertion, the CloBT curve is slightly lower but also peaks after the “Stress” phase and during “+Clothing” (Supplemental information 1.9a).

Similarly, CloBH showed a distinct rise during the “Stress” phase, with further increases frequently observed after the addition of clothing (Supplemental information 1.9b). The participants’ humidity levels peaked during “+Clothing” and remained elevated until the “Cool” phase began. Although individual variation was present, the general pattern showed a humidity build-up during stress exposure and partial recovery during the cool-down period. The observed pattern was more uniform across the three different stressor exposures compared to the CloBT response.

### Prediction of stress source

The importance of physiological and environmental factors used to classify stress varied depending on the type of stress—whether it was caused by temperature, humidity, or physical exertion. The full patterns are illustrated, depicting the variable importance per outcome class (Figure 2a-c.). Notably, the top 10 variables for each stress source revealed different dominance patterns: temperature stress predictors were largely environmental; exertion stress relied on a physiological mix (LSR, HR/SDNN, CBT); and humidity stress incorporated a balance of LSR, CloBT/CloBH, and HR. The difference in importance between the top 10 predictors and the rest is greater for ambient temperature during visits than for exertion and humidity.

**Figure 2.**
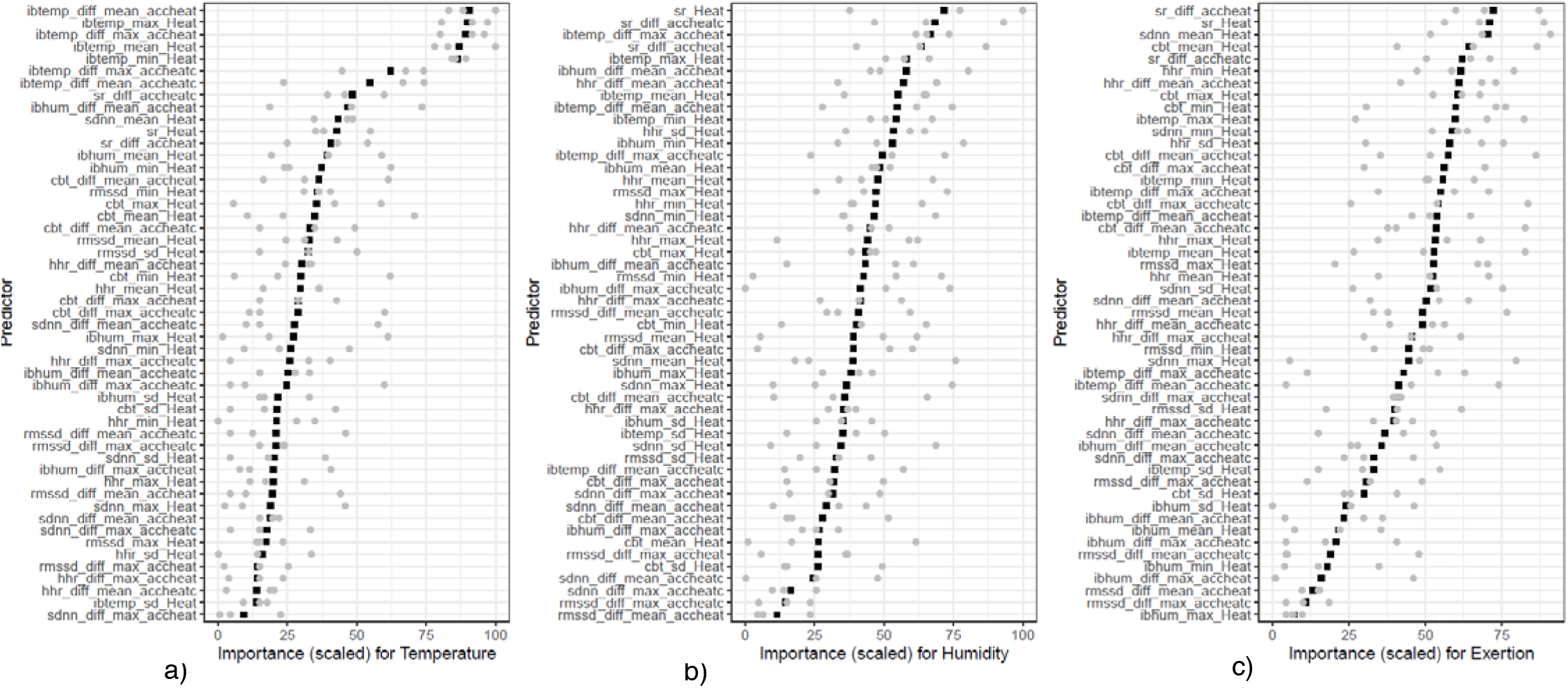
Predictors to identify heat stress sources. a) Variable importance for prediction of temperature stress. b) Variable importance for prediction of humidity stress. c) Variable importance for prediction of exertion stress. Top predictors identified by the Random Forest Model. Cbt indicates Core body temperature; hr indicates Heart rate, ibhum_: Close body relative humidity; ibtemp_: Close body temperature; rmssd indicates Root mean square of successive differences; sdnn indicates Standard deviation of nn intervals; and sr indicates local sweat rate. Statistical feature (middle): diff indicates difference between two phases; max indicates maximum value; mean indicates mean value; and min_indicates minimum value; sd_indicates standard deviation. Phase comparison or time window (suffix): Heat indicates stress phase; accheat indicates difference between the acclimatization and stress phase; and accheatc indicates difference of all three phases including acclimatization, stress and cool down.

The Random Forest model identified three temperature-related features from close body environment sensing as important predictors for detecting exposure to increased ambient temperature (Figure 2a). The most important is the mean difference in CloBT between the acclimatization phase and the heat phase (ibtemp_diff_mean_accheat), the second is the maximum CloBT recorded during the heat stress phase (ibtemp_max_Heat), and the third is the maximal difference in CloBT between the acclimatization phase and the heat phase (ibtemp_diff_max_accheat). Among all predictors assessed with the temperature stressor, close body conditions were consistently the most dominant, emphasizing the device’s potential in detecting exposure to ambient temperature.

For the humidity stressor, the Random Forest model identified absolute LSR during the heat stress phase (sr_Heat) and the change in LSR from acclimatization to the heat phase (sr_diff_accheatc) as the most indicative predictors (Figure 2b). The third most important variable was the maximum difference in CloBT between the “acclimatization” phase and the heat phase (ibtemp_diff_max_accheat), which was also among the predictors in the temperature model mentioned before. These three indicators contribute notably to the model’s ability to identify elevated humidity conditions in the study setting. In addition to LSR, several temperature-related features derived from close body environment are also ranked highly, including maximum and mean CloBT values across all phases. Compared to other physiological signals, LSR and CloBT consistently showed the greatest predictive importance, while HR and CBT contributed less to classification under humidity stress.

Lastly, the Random Forest model was computed for the exertion stressor (Figure 2c). In this case of exposure to exertion, a combination of physiological variables contributed to the model’s performance, including LSR, HR, CBT, and HRV metrics such as SDNN. The three most influential predictors for identifying exertion stress were: the change in LSR from the “acclimatization” to the heat phase (sr_diff_accheat), the absolute LSR during the heat “Stress” phase (sr_Heat), and the mean SDNN during the heat “Stress” phase (sdnn_mean_Heat). The first two were based on continuous plaster-based sweat rate measurements, while the third was derived from HRV data.

## Discussion

This study explored the potential of digital biomarkers for comprehensive health monitoring that includes physiological and environmental factors during exposure to heat. It was shown that different physiological and environmental signals, obtainable through wearable technologies such as smartwatches and sweat patches, can provide valuable health insights for each source of heat stress. Our results show that a Random Forest model could classify different heat stressor types – ambient temperature, elevated relative humidity, or physical exertion – with high accuracy (94.7%). This underscores the feasibility of data-driven heat stress differentiation in a controlled lab setting. Importantly, the variable contributing most to the model’s success varied by stressor: CloBT was most indicative of ambient temperature, LSR and CloBH were most relevant for relative humidity stress, and exertional stress was best identified by a combination of LSR, CBT, and HR features (including, particularly, SDNN, the standard deviation of normal-to-normal (NN) intervals, which reflects overall HRV over time). These findings provide a strong foundation for the potential value of a multimodal digital health sensing approach during heat exposure, when integrating both physiological responses and external environmental factors at the same time, thus allowing us to understand the human physiology in its contextual environment. With our study, we provide evidence that integrating environmental and physiological data might offer added value for assessing and ultimately managing people during heat exposure such as discussed by Jay et al.^11^.

Our approach is backed up by earlier research showing that physiological strain indices often predict heat-related illness more accurately than environmental indicators alone^4^. Wearable devices can support this approach by enabling long-term, individualized monitoring across various daily activities in real life^13^, generating valuable physiological data beyond the snapshot provided by traditional short-term assessments^14^. This is consistent with findings by Kwaro et al.^15^, who also demonstrated that combining physiological and environmental data enhances the accuracy of heat strain detection in field settings. It reflects a growing consensus that effective health monitoring in heat must integrate person-centered monitoring that goes beyond environmental metrics alone, incorporating physiological data to more accurately reflect individual responses to heat^16^.

Wearable sensors enable data collection beyond the laboratory, offering a more comprehensive view of human function in everyday settings. However, as Hafer et al.^21^ note, interpreting such real-world data requires careful attention to contextual factors, such as environment and activity patterns, that can significantly influence sensor outputs. Without this context, there is a risk of potentially misinterpreting variability as physiological changes. Concerning heat strain monitoring, this challenge is particularly relevant, as individual responses to heat are shaped not only by physiological parameters but also by external conditions in a complex interplay. This study demonstrates that integrating contextual data, such as the CloBT, with wearable-derived vital signs may enhance the assessment of health status under real-world heat exposure. In this regard, the case of the participant with the lowest HRV reactivity of all, who reported to be under major emotional stress due to the recent loss of a close family member, underscores the importance of interpreting an individual within their biopsychosocial context. Building on the importance of contextualizing wearable data, our findings illustrate how personalized heat strain monitoring might be effectively applied across both vulnerable and occupational populations. Among elderly individuals, particularly during extreme heat events, distinguishing between febrile responses due to infection and hyperthermia caused by environmental heat exposure remains a clinical challenge. Wearable devices that track vital signs such as CBT, HR, SR, or skin conductance, combined with contextual metrics like ambient temperature, indoor/outdoor status, and recent activity, could help resolve this ambiguity by providing individualized, real-time feedback. Furthermore, these systems could function as closed-loop alert or hydration tools, guiding users or caregivers to initiate timely cooling strategies or seek medical attention^10^.

A further dimension of individual variability in heat response is the degree of heat acclimatization, which remains difficult to quantify using current methods and recommendations. Although guidelines such as those from CDC/NIOSH (Acclimatization | Heat | CDC) recommend gradual exposure protocols for new and returning workers, they are not supported by individualized physiological data and lack objective markers of acclimatization status and are based on expert opinion evidence. Current assessments rely on indirect markers such as exposure duration or standardized heat response tests, yet these are often impractical outside laboratory settings and fail to capture continuous, personalized adaptation over time. This methodological heterogeneity limits the development of individualized heat management strategies and highlights the need for wearable multimodal systems capable of real-time acclimatization monitoring^22^. Multidimensional wearable sensing approaches enable continuous tracking of physiological adaptation over time and set these into context with varying environmental conditions, offering a more precise and actionable understanding of acclimatization. This multimodal strategy could support the development of evidence-based definitions of heat acclimatization, guide individualized prevention, and pre-/rehabilitation protocols.

To move beyond conventional vital parameter monitoring, the integration of diverse health modalities and dimensions allows for a more comprehensive understanding of individual heat vulnerability and its management. At the molecular level, recent work by Brasier et al.^12^ demonstrates that sweat-based biomarker candidates provide a potential non-invasive window into metabolic responses to heat stress, paving the way for biochemical personalization of heat exposure assessment. Further, the sweat rate is an emerging parameter in hydration monitoring^10^ and heat stress assessment, thus enabling the collection of digital biomarkers^23^. Complementing this, wearable ultrasound-based technologies enable continuous, cuffless monitoring of cardiovascular dynamics such as heart rate and blood pressure. Devices described by Hu et al.^24^ and clinically validated by Zhou et al.^25^ offer a high-fidelity measurement of cardiac function during real-world activity, providing dynamic insight into hemodynamic strain under heat exposure To account for environmental dimensions, wearables such as UV radiation sensors ^26^ allow individual-level qualification of solar exposure, an often-underappreciated factor in thermal load modeling. Recently, the European Environment Agency has emphasized that simultaneous exposure to heat and air pollution synergistically increases risks for cardiovascular and respiratory health, reinforcing the importance of incorporating pollution data into real-time decision-making systems^27^. Together, these multimodal and multidimensional wearable sensors have a major potential to develop meaningful and effective health monitoring tools in the heat. Further, biomarkers derived from continuous health sensing provide not only insights into health but also the ability to trigger just-in-time-adaptive interventions, such as hydration prompts, rest breaks, or environmental adjustments ^10,28^. A recent study demonstrated how such systems can deliver personalized alerts based on real-time heat strain data ^29^, highlighting their practical value in both everyday and occupational settings.

Translating the results from our study into practical, real-world applications reveals several interdisciplinary challenges that must be addressed. From a clinical standpoint, ensuring reliable, standardized, and interpretable output is essential, yet difficult due to individual variability in physiological responses, remaining technical limitations of current sensors, and a lack of policy consensus^8^. From the user’s perspective, wearable systems must be both comfortable and intuitive to ensure long-term adoption. At the same time, clinicians and healthcare providers require reliable and interpretable outputs that can meaningfully inform their decision-making and are reimbursable. Regulatory and industrial considerations add further complexity. Under the EU Medical Device Regulation (MDR 2017/745), such wearables, depending on the intended use, are classified as medical devices, triggering the need for rigorous clinical validation. Further, wearable devices contain software and increasingly often machine learning algorithms^17,18^. Thus, regulatory considerations must include both software as a medical device and the requirements of the AI (Artificial Intelligence) act. Further, reimbursement remains a significant hurdle: payers increasingly demand proof of cost-effectiveness, yet comprehensive health economic data for wearable technologies remains scarce. To better understand these multifaced challenges and identify pathways to integration, a Strengths-Weaknesses-Opportunities-Threats (SWOT) framework can offer valuable strategic insights. As highlighted in a recent analysis of smart patch technologies used by athletes^19^, such a framework can also help contextualize our findings and serve as a useful tool for assessing the integration of digital biomarkers’ findings into a wearable device. To further structure the evaluation of real-world integration, the Implementation Outcomes Framework by Proctor et al.^20^ provides an additional lens. It emphasizes key domains such as acceptability, adoption, feasibility, fidelity, and sustainability of health innovations within clinical and organizational contexts.

### Limitations

Despite promising explorative results, several limitations must be acknowledged. While our findings demonstrate the potential of a multimodal and multidimensional approach to monitor health during heat exposure, the study cohort, among others, was limited in size and thus did not represent the diversity of broader populations in terms of age, sex, fitness level, or environmental background. Expanding future studies to include more varied participant groups will be essential for developing inclusive and generalizable models. Additionally, the heat stress scenarios examined in this study were moderate, and highly controlled, potentially limiting the direct applicability of our findings to more extreme conditions, such as those experienced during heat waves. Moreover, our models were built on isolated heat stressors, which may not fully reflect the complexity of real-life situations where multiple stressors, such as dehydration, workload, or circadian variation, interact. Finally, the multimodal approach itself introduces practical challenges, as physiological signals such as sweat rate (a fluid-based measure), heart rate (electrical activity), and core body temperature (measured internally) each require distinct sensing technologies and integration methods. This complexity must be addressed to ensure wearable systems remain practical, accurate, reliable, and scalable for real-world applications.

Looking ahead, we see great potential for future research further validating the identified biomarker candidates with a focus on long-term deployment of wearable systems in complex real-world environments, particularly among underrepresented populations^15,29^. These efforts will enable the refinement of digital biomarkers, evaluation of behavioral responses to heat alerts, and support of their integration into clinical and public health. Finally, achieving this vision will require close collaboration between physiologists, engineers, data scientists, and public health experts. By bringing together technical innovation with real-world applications, more effective strategies can be developed to enhance the understanding and management of the health of individuals exposed to heat.

## Conclusion

Our study highlights the transformative potential of digital biomarkers in comprehensively monitoring health during heat exposure. By continuously assessing environmental and physiological data through wearable platforms such as CloBT/H, LSR, and HR, this approach supports real-time personalized health management conducted in the lived environment. This is particularly relevant in the context of increasing global temperature and more frequent heat-related health risks. Furthermore, targeted interventions across diverse settings, ranging from elderly care to occupational safety can be implemented. This would be a critical shift from static environmental thresholds to dynamic, individual heat impact management. By continuing to expand sensor capabilities and integrate them into user-responsive platforms, digital biomarkers may redefine how heat impacts on health are assessed, understood, and deterioration is prevented.

## Materials and Methods

### Study design and setting

This study was embedded into a larger single-center, prospective, cross-sectional, cross-over, observational exploratory study. The study design, including the randomized study visit sequence and participant recruitment, follows the framework outlined in Brasier et al.^12^. However, this manuscript focuses on the exploration of digital biomarker candidates towards digital health monitoring in the context of heat exposure. Healthy individuals between the ages of 18 and 40 years were recruited from Swiss universities. Inclusion and exclusion criteria, such as BMI below 30 kg/m^2^, heat-adapted, non-smokers, and limited regular physical activity (<4 hours per week), were consistent with the previous publication. Specific pre-visit conditions, hydration protocols, and other study guidelines were also followed as outlined in the earlier publication. The study was conducted at the Heat and Humidity Lab in Grenchen, Switzerland, between November 2022 and February 2023. Participants were instructed to avoid alcohol consumption and excessive physical activity for 24 hours prior to each visit, maintain proper hydration, and follow additional study-specific guidelines ^12^. Between the first and second visit, the CloBT and CloBH were measured during everyday life at home at the same time of day as the planned visits using a sensor (iButton® Hygrochron) worn around the neck. The mean CloBT and CloBH served as the personalized baseline conditions of the study. Visits 2 to 4 were conducted at the same time of day to adjust for chronobiological changes of body CBT over the day. Visits 2 to 4 consisted of four phases each, namely (i) the acclimatization phase (ii) the heat-stress phase (either increased ambient temperature (+ 10°C), increased relative humidity (+ 40%), or exertion (1 W/kg bodyweight)); (iii) the heat stress plus additional protective clothing (PC) phase (2 clo; +PC); and (iv) the cool-down. Each phase lasted 30 minutes. During visit 5, it was confirmed that all e-Celsius pills ingested during visits 2 to 4 had been fully excreted.

### Ethical considerations

The Ethical Committee of Northwestern Switzerland approved this observational study (EKNZ ID 2022-01325). All participants gave their informed consent prior to enrolment. The study was retrospectively registered at ClinicalTrials.gov (NCT05622188).

### Study objectives

This study aimed to examine the variability of physiological heat strain biomarkers, including HR, heart rate variability (SDDN and rMSSD), LSR, CloBT as well as CloBH, and CBT in a personalized as well as acclimatized setting. The objective was to identify distinct heat strain patterns associated with each type of heat stressor, to inform more person-centered health management strategies under elevated heat conditions.

### Sampling and measurements at the heat and humidity lab

As described in the previous publication on this study ^12^, HR was continuously monitored using a 3-lead Holter ECG (Bittium® Faros 360), while CBT was continuously monitored via an ingestible e-Celsius pill (BodyCAP®), taken 6 hours before each visit. AAs by gold-standard, LSR was measured using absorbent pads (TA; 3 M® Tegaderm+Pad – 6 ×10 cm (Sweat sampling pad 2.5 × 6 cm)), weighed before and after application with an electronic scale (precision of 0.001 g (Mettler and Toledo®, Switzerland)). The pads were applied to the anterior thorax, 3 cm below the mamilla, and assessed once per phase for four measurements per visit. The pads were stored in Eppendorf® tubes at −20°C for molecular analysis. Whole-body sweat loss was estimated by measuring body weight before and after each visit. Hydration status was checked using a handheld urine gravity device, and environmental conditions were monitored using a WBGT meter (TM-188D; Tenmars®).

### Sensing devices used for vital parameter measurements

#### iButton

Continuous measurement of the temperature and relative humidity of the close body environment was measured using small iButton temperature/humidity loggers (DS1923-F5, Hygrochron Temp/Humidity Logger, Maxim Integrated, USA). The loggers were worn around the neck attached to a comfortable, loose lanyard positioned above the T-shirt but beneath any jumper or outerwear (if worn). To establish a baseline for each study visit, only data points corresponding to the same time window as the visit were averaged across the previous seven days. As a result, the measured temperature and relative humidity were expected to represent the participant’s close body environment during everyday life during the time of day that matched the study visit.

### e-Celsius Performance system

CBT was continuously monitored using the e-Celsius pill (BodyCAP®; monitor firmware v6.1.0 and Performance Manager v1.4.2). The capsule is biocompatible, ingestible/non-digestible and complies with ISO 80601-2-56:2017 + AMD1: 2018. The participants were asked to ingest it 6h prior to their visit. The pill was then naturally excreted 2-5 days later, depending on the participants’ digestive system.

### Heart rate device

HR was continuously measured by a 3-lead Holter ECG (Bittium® Faros 360; Bittium cardiac navigator software v1.5.6).

### Sweat rate device/patches

LSR was measured using technical absorbents (TA; 3 M® Tegaderm+Pad – 6 ×10 cm (Sweat sampling pad 2.5 × 6 cm) placed on the anterior thorax just before the acclimatization phase started. The localization was chosen for sensors worn with a chest-strap. However, sweat from primarily eccrine glands is collected, thus devices might be worn on the arms – more often used collection localizations. Sweat was collected from the first minute of the subsequent phases. Technical absorbents were weighed before (dry weight) and after the phases (wet weight) in a sealed bag/tube to determine the final sweat rate. The bags were stored in a freezer at −20°C until lab-based sweat analysis was conducted.

### Statistical analysis

#### Descriptives

The flow of participants through the study was summarized by a CONSORT-like diagram depicting the number screened, seen at baseline visit, assigned to each heat stress sequence, and completed or dropped out. The participant characteristics were summarized in both overall and per heat stress sequence using the mean (standard deviation) for numeric variables and count (percent) for categorical data. The number of missing values was also reported in the participant’s characteristic tables. The laboratory environment was summarized by the mean (standard deviation) of the wet bulb globe temperature, ambient temperature, and humidity for each heat stress and phase combination.

Stress response measures (eCelsius; CBT, Holter monitor; HR, plaster-based patches; LSR) were summarized using means and standard deviations for each combination of stressor type (exertion, humidity, temperature) and phase (acclimatization, stress, stress+clothing).

For each participant, physiological and environmental data were continuously recorded throughout the 120-minute session and organized into time series plots. These plots were structured by phase (acclimatization, stress, stress with clothing, and cool-down), and on top have markers (lines) indicating the assigned heat stressor.

### Secondary analysis

This secondary analysis was meant to explore the feasibility of using heat stress response and environmental data to detect stress sources (temperature, humidity, exertion). A Random Forest model was built on 55 predictors derived from the last five minutes of the acclimatization, stress, and Stress+Cl phases. These predictors included means, standard deviations, minimums, maximums, and differences in means between the acclimatization phase and the stress or Stress+Cl phases. Data was collected from three sources: CBT from the e-Celsius pill, HR and HRV from the Holter monitor, and CloBT and CloBH from iButton sensors.

A Random Forest model was built to classify the stress source a participant was subjected to at a visit using predictors derived from the eCelsius, Holter monitor, and iButton the data contained multiple visits per subject, expected to be more like each other than to other participants. This was addressed by taking a random sample of the data, limited to one visit per subject. Repeating this process three times without replacement resulted in three datasets with independent observations. For each dataset, a Random Forest model was built using a leave-one-out cross-validation scheme to perform grid search tuning. The Model accuracy was averaged across leave-one-out folds and over the three datasets, yielding an estimated accuracy of 94.7% (95% CI: 81.6% - 100%).

## Data Availability

All data produced in the present study are available upon reasonable request to the authors.

## Acknowledgment

The authors would like to acknowledge support in the statistical analysis by Michael Prummer and Kelly Reeve from NEXUS Personalized Health, at ETH Zurich.

